# COVID-19: more than “a little flu”? Insights from the Swiss hospital-based surveillance of Influenza and COVID-19

**DOI:** 10.1101/2020.11.17.20233080

**Authors:** Georg Marcus Fröhlich, Marlieke E. A. De Kraker, Mohammed Abbas, Olivia Keiser, Amaury Thiabaud, Maroussia Roulens, Alexia Cusini, Domenica Flury, Peter W. Schreiber, Michael Buettcher, Natascia Corti, Danielle Vuichard-Gysin, Nicolas Troillet, Julien Sauser, Roman Gaudenz, Lauro Damonti, Carlo Balmelli, Anne Iten, Andreas Widmer, Stephan Harbarth, Rami Sommerstein

## Abstract

**Background:** Coronavirus disease 19 (COVID-19) has frequently been colloquially compared to the seasonal influenza, but comparisons based on empirical data are scarce.

**Aims:** To compare in-hospital outcomes for patients admitted with community-acquired COVID-19 to patients with community-acquired influenza in Switzerland.

**Methods:** Patients >18 years, who were admitted with PCR proven COVID-19 or influenza A/B infection to 14 participating Swiss hospitals were included in a prospective surveillance. Primary and secondary outcomes were the in-hospital mortality and intensive care unit (ICU) admission between influenza and COVID-19 patients. We used Cox regression (cause-specific models, and Fine & Gray subdistribution) to account for time-dependency and competing events with inverse probability weighting to account for confounders.

**Results:** In 2020, 2843 patients with COVID-19 were included from 14 centers and in years 2018 to 2020, 1361 patients with influenza were recruited in 7 centers. Patients with COVID-19 were predominantly male (n=1722, 61% vs. 666 influenza patients, 48%, p<0.001) and were younger than influenza patients (median 67 years IQR 54-78 vs. median 74 years IQR 61-84, p<0.001). 363 patients (12.8%) died in-hospital with COVID-19 versus 61 (4.4%) patients with influenza (p<0.001). The final, adjusted subdistribution Hazard Ratio for mortality was 3.01 (95% CI 2.22-4.09, p<0.001) for COVID-19 compared to influenza, and 2.44 (95% CI, 2.00-3.00, p<0.001) for ICU admission.

**Conclusion:** Even in a national healthcare system with sufficient human and financial resources, community-acquired COVID-19 was associated with worse outcomes compared to community-acquired influenza, as the hazards of in-hospital death and ICU admission were ∼3-fold higher.

## Introduction

The clinical impact of the ongoing COVID-19 pandemic is frequently compared to the seasonal influenza^1^. This may be due to its pattern to spread worldwide, but also because of a similar initial clinical presentation^2^. It is difficult to distinguish an infection with SARS-CoV-2 from an influenza A/B infection based on the clinical presentation. The most common symptoms are cough, fever, dyspnea or headache for both infections.

However, COVID-19 appears deleterious in many more aspects than influenza^3^; intensive care units reaching their limits, reduced number of elective procedures due to limited resources, national lockdowns to slow down the spread, and an unprecedented economic depression.

It is estimated, that up to 5000 patients with influenza A/B infection need hospitalization and approximately 1500 patients die each year during the influenza season in Switzerland^4, 5^. In comparison, COVID-19 has already caused more than 54.000 infections, 5400 hospitalisations and 1800 deaths in Switzerland until 2nd October 2020^6^.

While influenza is a well investigated infection with clear preventive and treatment strategies, including vaccination and anti-viral drugs, these strategies have not yet been defined for COVID-19. Novel treatment strategies with dexamethasone or remdesivir have been investigated, but are still in the process of approval^7,8^. At the same time, 195 SARS-CoV-2019 projects have been registered to develop a dedicated SARS-CoV-2019 vaccine ^9^. Of these, 9 candidates are currently evaluated in phase III trials^9^.

So far, only a few, small scale, retrospective studies directly compared hospital outcomes of COVID-19 and influenza infections^10, 11^. These studies demonstrated, that in COVID-19 as well as in patients with influenza infection, many patients have underlying co-morbidities that may impact on the course of disease independently^10, 11^. Due to the small sample size of these previous studies, it has been difficult to draw meaningful conclusions. However, the comparison of COVID-19 and seasonal influenza is important from a public health perspective. Therefore, we aimed to investigate differences in mortality and ICU admission among patients hospitalized for COVID-19 and influenza in Switzerland.

## Methods

### Hospital-based surveillance of COVID-19/Influenza cases in Switzerland – database characteristics and study population

All data are derived from the prospective database of the *Hospital-based surveillance of COVID-19/Influenza cases in Switzerland*. The database was originally initiated to collect data for all hospitalized influenza cases during the Influenza season 2018/19 by the Institute of Global Health at the University of Geneva, Switzerland. In total, 14 Swiss based hospitals, including all Swiss university hospitals contributed data. Seven centers collected data for all patients who were hospitalized with influenza A/B (start date: 28 October 2018) and 14 centers (including 5 overlaps) for SARS-CoV-2 (start date: 19 February 2020).

A standardized, central, REDCap hosted electronic questionnaire for data collection was set up for data entry in the participating centers^12, 13^.

Hospital data included patient demographics, main diagnosis, non-invasive or invasive ventilation, cardiovascular, pulmonary or neurologic complications, hospital and ICU admission and discharge date, as well as co-morbidities or death^12, 13^. Inclusion required a positive PCR based test obtained from any respiratory sample.

Only data for completed hospitalizations of patients >18 years up to July 24th 2020 were included for the analyses. Potential hospital-acquired infections (with PCR diagnosis obtained later than 2 days after admission) were excluded to focus on patients with influenza or COVID-19 present on admission.

The study was approved by the Ethics Committee of the Canton of Geneva, Switzerland. (CCER 2018-00577, 2020-00827). Data collection was also approved by all local Ethics Committees.

#### Outcome measures

The primary outcome measure was defined as *in-hospital all-cause mortality*. Secondary outcome was admission to the Intensive Care Unit (ICU). Further exploratory end-points included cardiovascular, pulmonary, renal or neurologic complications during hospitalization, length of stay and antibiotic treatment.

#### Statistical analysis

For the primary and secondary outcome analyses we used cause specific cox hazard models for the outcome of interest, while accounting for competing risks (hospital discharge for hospital mortality; discharge and death before ICU admission for ICU admission) to prevent overestimation of the impact of COVID-19^14^. Patients that stayed more than 30 days were right-censored in all models. To adjust for differences in baseline characteristics, inverse probability weighting was applied using clinical and epidemiological factors present at admission significantly associated with influenza versus COVID-19, as determined by univariate logistic regression models^15^. We applied truncation at 1^st^/99^th^ percentile. The method and results of the validation of the weights is provided in Supplementary Table 1. In a second step, subdistribution hazard analysis using the Fine & Gray model were applied to determine the cumulative risk of death or ICU admission associated with COVID-19 versus influenza. The main analyses were adjusted for gender, age, and university hospital as treatment center (available for all cases). In a subgroup analysis we adjusted in addition for body weight and the presence of comorbidities. Since this information was not systematically collected by all centers we included only patients/centers where this info was available (n=2634; 1731 for COVID-19 and 903 for Influenza). There, we also adjusted for weight and presence of comorbidities (cardiovascular, asthma, respiratory, hemato-immunological, renal and neurological; Suppl. Table 2). Further subgroup analysis focused on i) “in-hospital transfers” to the ICU, excluding direct admission to the ICU from the community and ii) community transfers, excluding patients admitted from long term care facilities (LTCF), as during the COVID-19 pandemic fewer LCTF cases may have been transferred to hospital to prevent overburdening of hospital capacities. As the date of outcome was only recorded for mortality and ICU admission, we only calculated the crude incidence for all other outcomes. All analyses were performed in “R (R Core Team, Version 4.02)”, using packages *survival, ipw, CausalGAM* and *cmprsk*^16^. Two-tailed tests were performed and p-values <0.05 were considered statistically significant.

## Results

### Baseline characteristics

In total, 4441 patients were available from the two surveillance databases, of which 217 patients had to be excluded (170 for COVID-19 and 47 for Influenza), because hospital admission or discharge date were missing.

In total, 2843 patients with a confirmed diagnosis of community-acquired COVID-19 were included from 14 acute care hospitals all over Switzerland between 19th February 2020 and 22nd July 2020, including all 5 Swiss university hospitals. This represents 50% of all reported hospitalizations in Switzerland^17^ (4569 hospitalizations as of 3^rd^ September 2020). Between 26th October 2018 and 26th March 2020 1331 patients (96.4%) with influenza A and 50 patients (3.6%) with influenza B were included from the influenza database from 7 centers, including 3 university hospitals. The majority of patients with COVID-19 (2476/2843, 87%) as well as those with influenza A/B (1220/1381, 97%) were directly referred to one of the participating hospitals. Several patients were transferred from local district hospitals to specialized facilities (4.9 % of COVID-19 patients and 3.3 % of Influenza patients), 4.8% (n=137) of the COVID-19 patients were transferred from a LTCF compared to 5.2% (n=72) of the influenza patients.

On average, patients with COVID-19 were younger, than patients with influenza A/B infection (median 67 years [IQR 54-78] versus 74 years [IQR 61-84], p<0.001) and were predominantly male (n=1722, 61% vs. 666 influenza patients, 48%, p<0.001), (Table 1).

**Table 1.** Baseline characteristics of COVID-19 and influenza patients in the Swiss surveillance database.

| Characteristics | SARS-CoV-2<br>(n=2843) | Influenza A/B<br>(n=1381) | p-value |
| --- | --- | --- | --- |
| Age (years), median (IQR) | 67 (54-78) | 74 (61-84) | <0.001 |
| Female sex, n (%) | 1121 (39.4) | 715 (51.8) | <0.001 |
| Admission to university hospital, n (%) | 1597 (56.2) | 935 (67.7) | <0.001 |
| BMI (kg/m<sup>2</sup>), median (IQR) | 26.7 (23.7, 30.5)<sup>+</sup> | 25.2 (21.8, 29.4)<sup>++</sup> | <0.001 |
| Comorbidities, n (%) |  |  |  |
| Diabetes mellitus | 522 ( 26.9) * | 291 (28.2) ** | 0.578 |
| Chronic cardiovascular disease | 737 ( 38.0) * | 473 (45.9) ** | <0.001 |
| Renal impairment | 374 ( 19.3) * | 243 (23.6) ** | 0.011 |
| Chronic pulmonary disease | 413 ( 21.3) * | 322 (31.2) ** | <0.001 |
| Chronic neurologic impairment | 276 ( 14.2) * | 190 (18.4) ** | 0.002 |
| Haematological disorder | 39 ( 2.0) * | 160 (15.5) ** | <0.001 |
| Chronic liver disease | 114 ( 5.9) * | 57 ( 5.5) ** | 0.873 |

Missing values n (%): <sup>+</sup>856 (30%), <sup>++</sup>518 (38%), <sup>\*</sup>904 (32%), <sup>\*\*</sup>350 (25%), as reporting of BMI/comorbidities was not mandatory and omitted by some centers. IQR, interquartile range; BMI, body mass index

### Outcomes

Table 2 gives an overview of the crude clinical outcomes for patients with COVID-19 or influenza A/B. Crude hospital mortality among COVID-19 patients was almost three times higher than for influenza A/B patients (363, 12.8% in COVID-19 patients and 61 patients, 4.4% with influenza A/B). ICU admission was also more frequent among COVID-19 patients (483, 19.4%) compared to influenza patients (129, 9.8%). Similarly, invasive ventilation was more common in COVID-19 patients (394, 13.9%) compared to influenza patients (66, 4.8%, p<0.001).

**Table 2.** Unadjusted crude outcomes for patients with microbiologically confirmed community-acquired COVID-19 or influenza infections.

|  | **SARS-CoV-2**<br>**(n=2843)** | **Influenza A/B**<br>**(n=1381)** | **p-value** |
| --- | --- | --- | --- |
| In-hospital deaths, n (%) | 363 (12.8) | 61 ( 4.4) | <0.001 |
| Admission to the ICU, n (%) | 483 (19.4) | 129 ( 9.8) | <0.001 |
| Invasive ventilation, n (%) | 394 (13.9) | 66 ( 4.8) | <0.001 |
| Respiratory complications, n (%) | 2082 (93.6)* | 744 (83.9)** | <0.001 |
| Cardiac disease, n (%) | 556 (25.0) * | 212 (23.9)** | 0.291 |
| Neurologic impairment, n (%) | 394 (17.7)* | 82 ( 9.2)** | <0.001 |
| Kidney failure, n (%) | 601 (27.0)* | 149 (16.8)** | <0.001 |
| Length of hospital stay, median (IQR) | 8 (5-16) | 7 (4-13) | <0.001 |
| Any antibiotic treatment, n (%) | 1539 (61.7) <sup>+</sup> | 862 (65.7) <sup>++</sup> | <0.001 |

Missing values n (%): \*618 (22%), \*\*494 (36%), <sup>+</sup>349 (12), <sup>++</sup>69 (5), as reporting of these complications was not mandatory and omitted by some centers. ICU, intensive care unit; IQR, interquartile range.

The results of the cause-specific cox hazards ratios are available in Table 3. The adjusted subdistribution Hazards Ratio (sdHR) for in-hospital mortality was 3.01 (95% CI 2.22-4.09, p<0.001) for COVID-19 versus influenza, and 2.44 (95% CI, 2.00-3.00, p<0.001) for ICU admission (selected variables and balance of weights in Supplementary Table 1). The cumulative incidence plots of the primary and secondary outcomes are shown in Figures 1 & 2: Most patients were discharged alive within a week, whereby discharge rates were higher for influenza. Most deaths occurred after the first week, and steadily increased up to two weeks after admission, again rates were higher for COVID-19. ICU admission mostly occurred early during stay (Figure 2).

**Table 3.** **Adjusted hazard ratios of primary and secondary outcomes (COVID-19 vs. influenza) by weighted cause-specific (CS) Cox models. Patients with hospital duration >30 days were right-censored at day 30.**

|  | **CS Hazard**<br>**ratio** | **95%**<br>**Confidence**<br>**interval** | **p-value** |
| --- | --- | --- | --- |
| Primary outcome analysis |  |  |  |
| Hospital mortality | 2.58 | 1.90-3.50 | <0.001 |
| Hospital discharge | 0.78 | 0.72-0.84 | <0.001 |
| Secondary outcome analysis |  |  |  |
| ICU admission | 2.46 | 2.00-3.01 | <0.001 |
| Death before ICU admission | 3.93 | 2.61-5.95 | <0.001 |
| Discharge alive before ICU admission | 0.95 | 0.87-1.03 | 0.181 |

**Figure 1.**
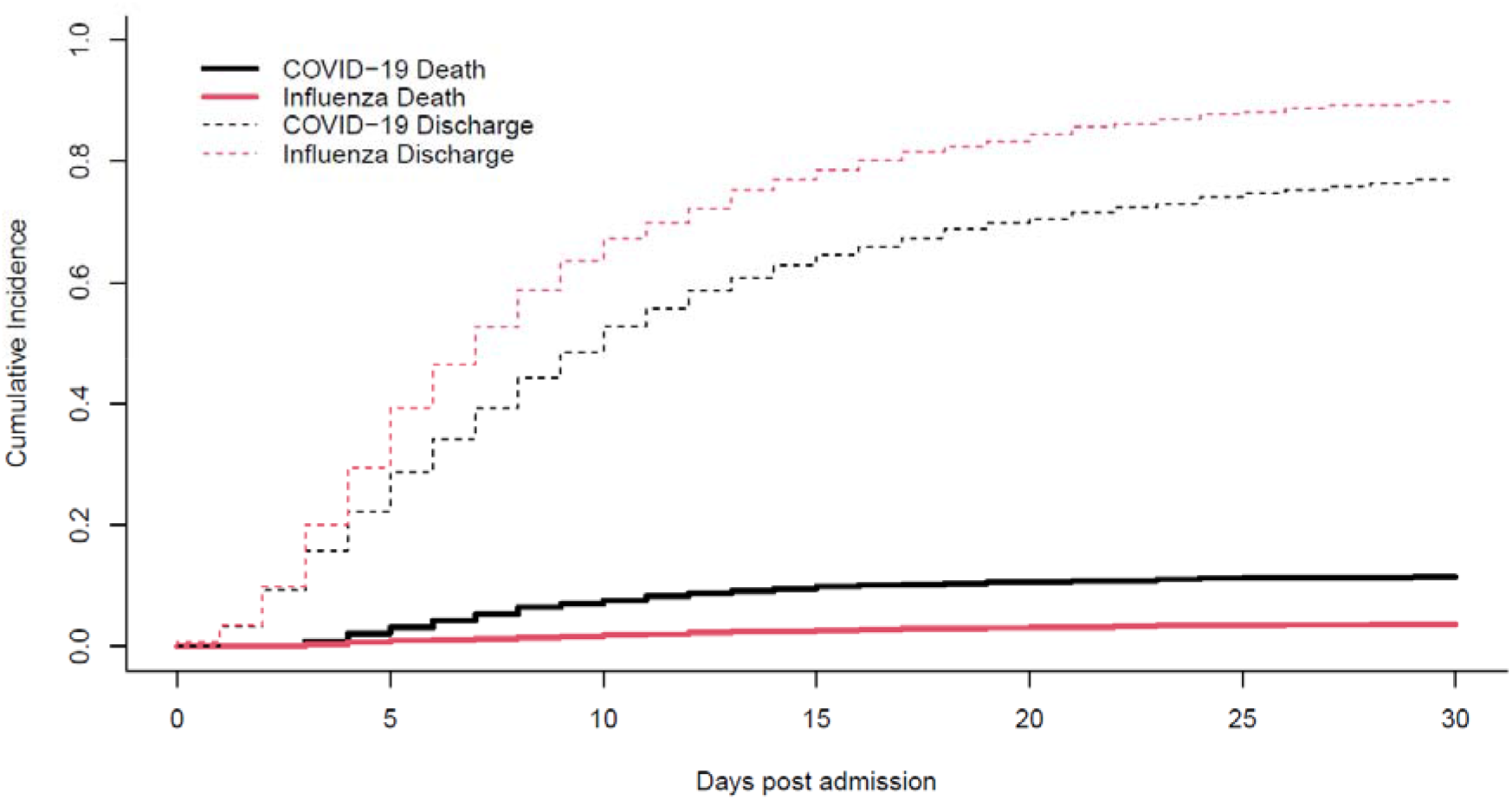
**Cumulative incidence plot. Mortality with discharge as competing risk, by disease status (COVID-19 versus influenza)**

**Figure 2.**
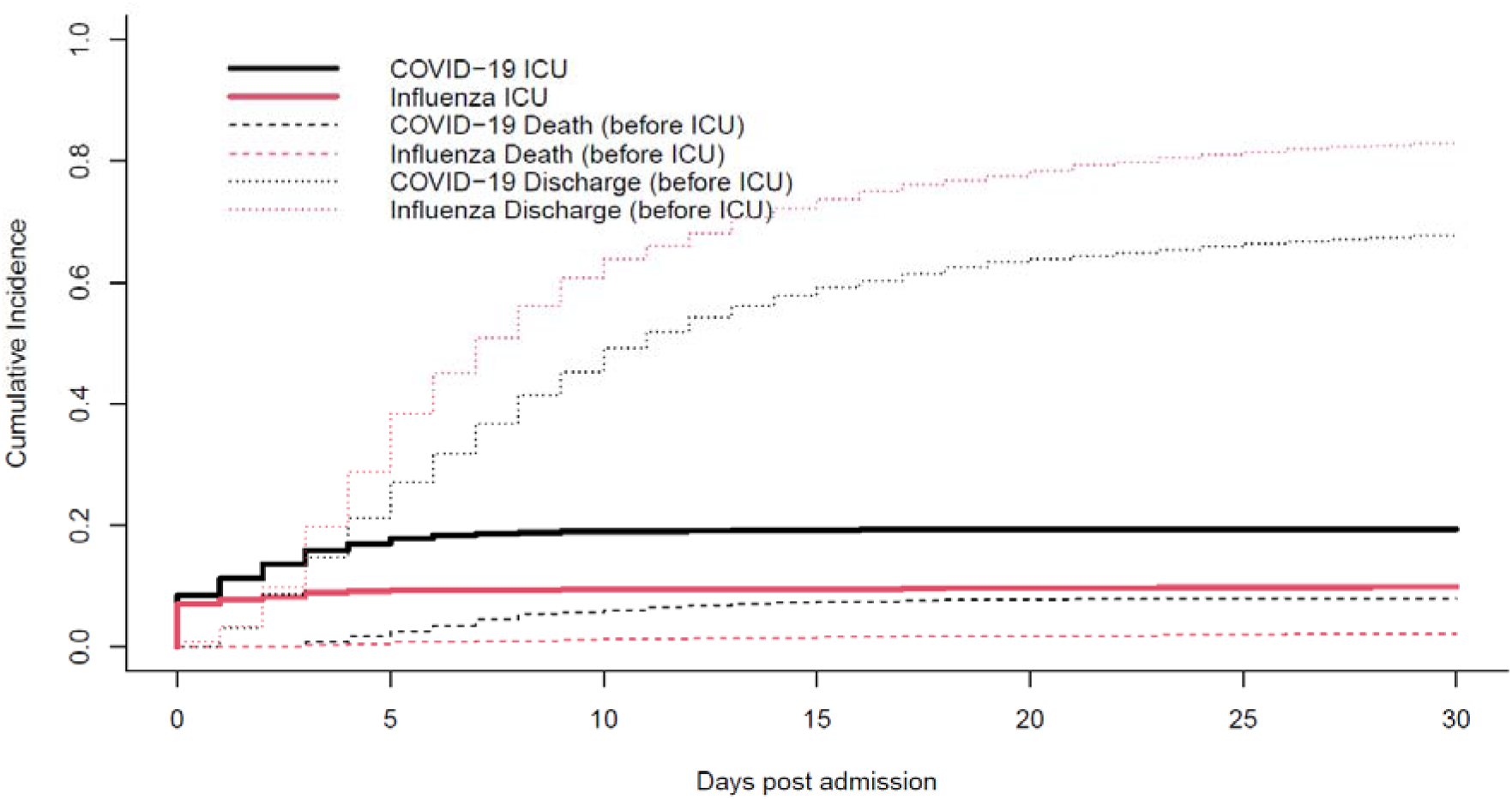
**Cumulative incidence plot. ICU admission with discharge and death before ICU admission as competing risk, by disease status (COVID-19 versus influenza)**

The adjusted sdHR for mortality within the 2634 complete cases was 3.14 (95% CI 2.11-4.68, p<0.001; balance of weights for confounders are shown in Supplementary Table 2). The adjusted sdHR for ICU admission, excluding the n=302 patients directly being admitted to the ICU on the day of hospitalization was 5.05 (95% CI, 3.54-7.20, p<0.001; cumulative incidence plot in Suppl. Fig 1). The adjusted sdHR for mortality in the subgroup, excluding n=209 direct transfers from LTCF was 3.10 (95% CI, 2.23-4.32; p<0.001).

## Discussion

This study directly compared the outcomes of patients hospitalized for influenza A/B or COVID-19 in a prospective nation-wide Swiss surveillance database. After taken account of competing events as well as imbalances between patient groups, community-acquired COVID-19 was associated with a 3-fold increased cumulative risk to die within hospital, compared to an influenza A/B infection. This increase was associated with a higher daily mortality risk as well as an increased length of stay in hospital for COVID-19 patients. Similarly, COVID-19 patients had a more than two-fold increased risk of being transferred to the ICU than influenza patients.

The COVID-19 pandemic poses a tremendous medical, socio-economic and also financial burden for many countries. In the absence of a curative treatment or vaccination, it is impossible to offer easy solutions. At the same time, COVID-19 is sometimes downplayed to be only a „little flu”^1, 18, 19^. However, the virus characteristics, as well as the course of disease in COVID-19 appear to be more serious than for influenza in several aspects: SARS-CoV-2 appears to be more contagious with a reproductive number of 2-2.5 and is contagious even in patients without symptoms or before symptoms appear^2^. The WHO reports, that 80% of COVID-19 cases are mild, 15% might need hospitalization and 5% of hospitalized patients require ventilation^2^. In comparison, a study from Germany including 34.493 patients with influenza infections demonstrated, that 3.7% of patients needed hospitalization, and of these 5% received invasive ventilation^20^. In our study, 13.9% of the hospitalized COVID-19 patients needed mechanical ventilation, compared to 4.7% of the influenza patients. These numbers underline the clinical importance of SARS-CoV-2 infections.

In our study, patients with COVID-19 were predominantly male and had a median age of 67 years, while influenza patients were predominantly female and older (median age 74 years). Therefore, the *years of potential life lost* are likely to be higher in COVID-19, which is underlined by the fact that COVID-19 patients were less likely to have significant co-morbidities than influenza patients. At the same time, all their clinical outcomes were worse than those for influenza patients, which persisted even after adjustment for possible confounders. The WHO estimates infection fatality rates of 3-4% for COVID-19, while infection fatality rates are usually much lower in the range of 0.1% for seasonal influenza infections^2^. In our Swiss hospital surveillance, the cumulative incidence of mortality of hospitalized COVID-19 patients at day 30 was 11.4% compared to 3.6% in hospitalized influenza patients. Previous studies reported mortality rates of hospitalized COVID-19 patients in the range of 4% to 28%^21, 22^. In addition, up to 2^nd^ October 2020 already 4890 COVID-19 cases have been hospitalized in Switzerland, which has resulted in more than 1800 associated deaths, which is higher than the total number of influenza-associated deaths in previous influenza seasons ^5, 23^.

In the presented Swiss cohort, the cumulative incidence of COVID-19 patients with ICU admission up to day 30 was 19.4%, compared to 9.9% in the influenza group. Interestingly, influenza patients who needed intensive care, were transferred to ICU early after hospital admission. In contrast, the proportion of ICU transfers in COVID-19 patients increased steadily up to 5 days after hospital admission (Figure 2).

Apart from respiratory complications, also neurologic impairment or kidney failure were more common in the COVID-19 cohort. It is unclear whether this finding might be explained by vasculitis and thromboembolic events, which are common in COVID-19^24^. Surprisingly, there was no difference in cardiovascular complications. While antibiotic therapy was more common in patients with influenza in this Swiss cohort, it remains unclear whether influenza is more often associated with pulmonary bacterial superinfection than COVID-19.

The present study has several limitations. The impact of various treatment strategies could not be analyzed in the presented Swiss cohort. It can be assumed that mortality for the included COVID-19 patients was relatively high, since the majority of the presented cases occurred early during the COVID-19 pandemic, and knowledge about the best supportive treatment strategies has slowly increased over time. For influenza treatment strategies have been more clearly defined, so the current findings may not apply to future COVID-19 cases.Of note, ∼1/3 of comorbidity data was missing, because it was not mandatory for the surveillance. As it was missing at random, specific centers did not report this data, we carried out a complete cases analysis for the primary outcome. The point-estimate of the complete case analysis fell within the 95% CI of the main analysis, indicating that absence of this data did not introduce a major bias.

Post-discharge follow-up was not available neither for influenza nor for COVID-19 patients, as such differences in long-term morbidity could not be discerned while it is assumed that these may have a higher incidence among COVID-19 cases. Finally, since no data was available for co-infection with influenza and SARS-CoV 2 this could not be considered. However, a study from Wuhan found that only 4% of COVID-19 patients had a co-infection with influenza, ^25^ and no difference in the course of disease was noted in comparison to COVID-19 alone^25^.

## Conclusion

In this prospective Swiss, multi-center hospital cohort, COVID-19 was associated with a 3-fold higher hazard for hospital mortality and for ICU transfer compared to seasonal influenza A/B infection. This indicates that among hospitalized patients, COVID-19 versus influenza has a larger excess mortality, and COVID-19 cannot be considered as a “little flu” in this cohort

## Data Availability

Swiss Covid-19/Influenza Hospital Surveillance data access is available from: https://www.unige.ch/medecine/hospital-covid/files/4015/9427/6937/CH_SUR_Multicentric_process_final.pdf

https://www.unige.ch/medecine/hospital-covid/

## Acknowledgments

We thank all participating centers for providing their surveillance data.

These data were collected in collaboration with the Swiss Federal Office of Public Health. With contributions of the Clinical Research Center, Geneva University Hospitals and Faculty of Medicine, Geneva. The authors want to especially thank Markus Schneemann, Department for General Medicine, Spitäler Schaffhausen, Promenadenstrasse 21, 8200 Schaffhausen, Switzerland; Laurence Senn, Hygiene, prevention et controle de l’infection, Centre Hospitalier Universitaire Vaudoise, Rue du Bugnon 21, CH-1011 Lausanne, Lisa Brockhaus, Department for General Medicine, Hôpital du Jura, Fbg des Capucins 30, 2800 Delémont, and Jonas Marschall, Bern University Hospital for acting as local principal investigators in the hospital-based surveillance.

## Author’s Role

Conception and Design: GF, RS, MdK, SH, AW, CB, MA. Acquisition of Data: AI, OK, AT, MR, AC, DF, PWS, MB, NC, DVG, NT, RG, LD, CB, AW, AW, SH

Statistics: MdK, RS, SH, JS, MA; Writing of first draft: GF, RS, MdK. Interpretation and Analysis: All authors; RS is guarantor of the paper. All authors have seen, critically revised and approved the manuscript.

## Data sharing statement

Swiss COVID-19/Influenza Hospital Surveillance data access is available from: https://www.unige.ch/medecine/hospital-covid/files/4015/9427/6937/CH_SUR_Multicentric_process_final.pdf

## Supplementary Materials

**Supplementary Table 1:** Balance check of inverse probability weights. We used balance.IPW from the CausalGAM package (Ref R): This function calculates weighted means of covariates and then examines the differences in the weighted means across influenza and COVID-19 cases as a diagnostic for covariate balance for inverse probability weighting. The standardized mean differences between all variables in the dataset are reported along with a z-statistics for these standardized differences, whereby z-statistics closer to 0 imply better univariate mean balance. The columns are (from left to right) the observed mean of the covariate among the COVID-19 patients, the observed mean of the covariate among the influenza patients, the weighted mean of the covariate among the COVID-19 patients, the weighted mean of the covariate among the influenza patients, the weighted mean difference, and the z-statistic for the difference. The formula (logistic regression with logit-link) used for calculation of weights was: Virus_type ∼ Age + Gender + Admission_to_University Hospital

|  | obs.mean.<br>C | obs.mean.<br>I | w.mean.C | w.mean.I | w.mean.di<br>ff | z |
| --- | --- | --- | --- | --- | --- | --- |
| Age | 65.11 | 70.41 | 66.76 | 66.43 | 0.33 | 2.252 |
| Gender (% female) | 0.39 | 0.52 | 0.44 | 0.45 | -0.01 | -3.717 |
| Admission to<br>University hospital<br>(%) | 0.56 | 0.68 | 0.60 | 0.60 | -0.00 | -0.689 |

**Supplementary Table 2:** Balance check of inverse probability weights for complete case analysis. The formula (logistic regression with logit-link) was: Virus_type ∼ Age + Gender + Admission_to_University Hospital + weight + com_cardiovascular + comorbidity_asthma + comorbidity _neuro_impair + comorbidity _renal + comorbidity _hemato_immuno The table contains the same information as explained above (Supplementary Table 1).

|  | obs.mean.Covid-19 | obs.mean.Influenza | w.mean.Covid-19 | w.mean.Influenza | w.mean.diff | z |
| --- | --- | --- | --- | --- | --- | --- |
| Age | 68.957 | 73.196 | 70.788 | 70.667 | 0.122 | 0. |
| Gender (female) | 0.406 | 0.494 | 0.439 | 0.445 | -0.006 | -0. |
| Admission to university hospital | 0.515 | 0.681 | 0.573 | 0.577 | -0.004 | -0. |
| Weight | 79.253 | 73.37 | 77.023 | 77.457 | -0.434 | -1. |
| Comorbidities (Yes/No) |  |  |  |  |  |  |
| Cardiovascular | 0.392 | 0.464 | 0.423 | 0.423 | 0 | 0. |
| Asthma | 0.091 | 0.134 | 0.108 | 0.109 | -0.001 | -0. |
| Neurological impairments | 0.139 | 0.171 | 0.149 | 0.16 | -0.011 | -3. |
| Renal | 0.201 | 0.252 | 0.228 | 0.223 | 0.005 | 0. |
| Hematological/immunological | 0.021 | 0.159 | 0.074 | 0.065 | 0.009 | 1. |

**Supplementary Figure 1.**
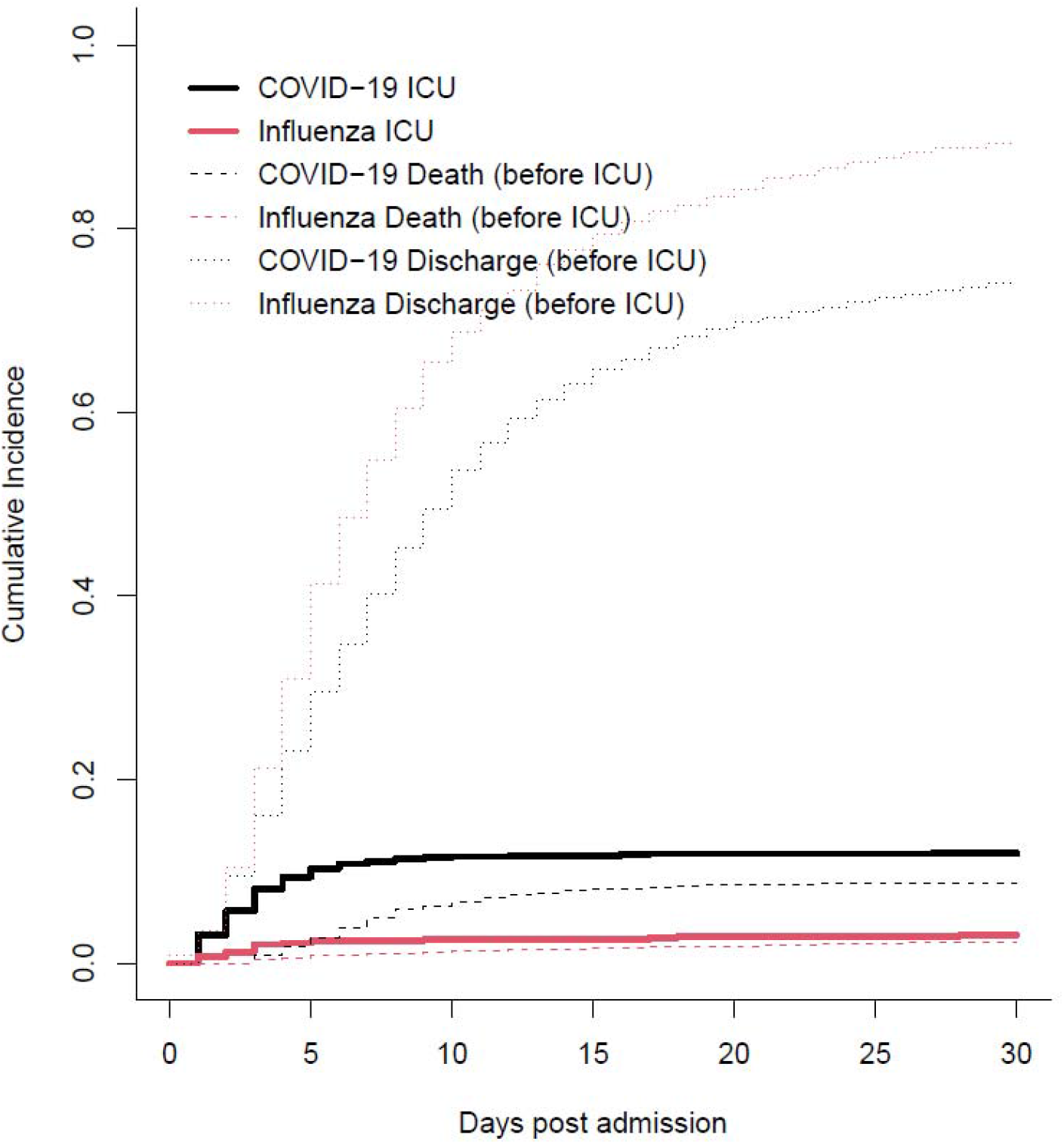
**Cumulative incidence plot. ICU admission with discharge and death before ICU admission as competing risk, by disease status (COVID-19 versus influenza); Patients directly admitted to the ICU from the community were excluded for this subgroup analysis**.

